# Impact of data labeling protocol on the quality of LGE-MRI atrial segmentation

**DOI:** 10.1101/2024.06.14.24308940

**Authors:** A.K. Berezhnoy, A.S. Kalinin, D.A. Parshin, A.G. Selivanov, A.G. Demin, A.G. Zubov, R.S. Shaidullina, A.A. Aitova, M. M. Slotvitsky, A.A. Kalemberg, V.S. Kirillova, V.A. Syrovnev, V.A. Tsvelaya

## Abstract

Atrial fibrillation affects up to 2% of the adult population in developed countries, and ablation as the main method of treatment leads to a high probability of recurrence. For such procedures, the approach of creating an in silico model of the patient’s atrium to be used for navigation during the catheter ablation procedure itself is extremely promising. In this case, the MRI data on which the model is based must be loaded into the system and segmented with high accuracy. This paper describes a new universal protocol for the segmentation of LGE MRI images. This protocol has been used to train state-of-the-art neural networks for automatic MRI segmentation. It is shown that the new data labeling protocol significantly improves the training quality of the network. Using this approach, it is possible to improve the quality of the reproduction of the patient’s atrial parameters and the performance of all related services. The presented protocol is also accompanied by a labeled image dataset. In the future, the data from such labels can be used for predictive modeling and the creation of digital twins of patients’ atria.

## Introduction

Atrial fibrillation (AF) currently affects approximately 2% of the adult population in developed countries, and within 20 years the prevalence may reach 60% in people over 60 years of age [1]. The main treatment for persistent AF is surgical ablation - destruction of the atrial myocardium in the arrhythmogenic zones. This leads to necrosis and apoptosis of cardiac tissue, which is eventually replaced by fibrous tissue. The latter does not conduct an electrical impulse, but in some cases may be an independent substrate for re-entry [2]. According to international clinical guidelines for the treatment of atrial fibrillation [3], isolation of the pulmonary vein ostia is always performed first. Other approaches have not shown significant efficacy in the treatment of AF and are not widely used [4]. It has been shown that more than 40% of patients have recurrent AF, which may require repeated surgical interventions [5, 6, 7] due to the extrapulmonary localization of the arrhythmogenic substrate. To address this problem, we are developing a model of the patient’s atrium that will automatically predict the optimal surgical protocol (the optimal pattern of damage caused) that will reduce the likelihood of the patient returning for reoperation. This approach has already shown great potential, for example in the work of Trayanova et al [8]. The authors were able to reduce the probability of recurrence by more than 30%. We expect better results because we take into account cellular interactions in the modeling, which has been shown [9] to make wave dynamics modeling more accurate. The goal of this work is to create a 3-dimensional model of a patient’s atrium based on their MRI data as part of the ablation correction system described.

An important part of such a system is the segmentation of MRI images of the patient’s atria with contrast (gadolinium): by looking for the atrial walls and the fibrotic foci themselves, it is possible to visualize the topology of the atrium and the fibrosis in the tissue for a correct prediction of the wave dynamics in it. The developed model itself has the potential to work with existing navigation systems for ablation, such as CARTO [10]. Automatic image segmentation today is usually performed using artificial intelligence (AI) methods. For this purpose, a dataset for training a neural network (NN) has to be assembled with manually labeled atrial MRI images, and the quality of the segmentation and its homogeneity can significantly affect the result of NN training. It has been shown [11] that the interpretation of images in different clinics (by different specialists) can be very different, which may ultimately lead to an incorrect diagnosis. Therefore, the main task in preparing the dataset to ensure its homogeneity was to develop a protocol for the atrial walls segmentation on MRI images; this is one of the main results described.

According to the developed protocol, a team of qualified cardiologists and cardiac surgeons manually labeled both open source and hand-collected images. The composition of the dataset is described in the results of the work. An important advantage over other available datasets is that several specialists participated in the labeling.

Based on these data, a neural network was created and trained to segment the atrial walls on MRI images. Experiments were conducted with different types of architectures and their combinations, and the most successful approach to image enhancement was selected. Separately, tools for atrial blood pool segmentation [12] and other pre-trained neural networks [13, 14] were considered as pre-processing. Using the labeled data, experiments were performed to segment the blood pool of the left atrium, the wall of the left atrium, and the left atrium completely on MRI images; for the atrial cavity, the dice on the test sample was 0.9, for segmentation of the atrial wall - 0.64.

## Materials and methods

### Data acquisition and Preprocessing

Two datasets were used. The first one: cDEMRIS (Cardiac Delayed Enhancement Segmentation Challenge) was taken from open sources [12]; only the images themselves, but not the atrial wall segmentation, are publicly available. The MRI images in this set were obtained from three centers: Utah School of Medicine, Beth Israel Deaconess Medical Center (BIDMC), and King’s College London Imaging Center (KCL-IM). The set includes high-resolution MRI images of the left atrium with contrast (used to visualize areas of fibrosis and scarring in the left atrial myocardium) for 30 patients - 30 images before and 30 images after ablation, for a total of 60 images. Each center provided 10 pre and post ablation images.

The second data set also consisted of delayed-contrast MR images of the left atrium. All patients with persistent AF before and after ablation underwent cardiac MRI on a 1.5 Tesla magnetic resonance imager (Magnetom Aria, Siemens, Germany). To evaluate structural changes in the left atrium (LA), an inversion recovery (IR) gradient sequence with fat signal suppression and an isotropic voxel of 1.25×1.25×2.5 mm reconstructed in 0.625×0.625×2. 5 mm (TR - repetition time - corresponding to the R-R interval (600 to 1100 ms); TE - echo time - 2.44 ms; magnetization vector deflection angle - 22 g; slice thickness - 2.44 ms; slice thickness - 2.5 Tl.; slice thickness - 2.5 mm; field of view - 30-35 cm; eight averages; 40-50 slices) [12]. This sequence was performed 15-20 minutes after intravenous bolus injection of gadolinium-based contrast agent (Gadovist) at a dose of 0.15 mmol/kg. The MRI study was performed under the conditions of respiratory synchronization against the background of free breathing of the patient and synchronization with ECG. Data acquisition was performed in the phase of atrial diastole during exhalation, which was determined by the position of the right diaphragmatic phrenic. The technical parameters of the images from all databases used in the work are presented in Table 1.

**Table 1.** The technical parameters of the MRI images from 2 databases used in the work: open database cDERMIS [12] and our new original database VMRC.

|  | cDEMRIS |  |  | New original database (VMRC-data) |
| --- | --- | --- | --- | --- |
| Dataset | U. Utah | BIDMC | KCL-IM | M. F. Vladimirsky |
|  |  |  |  | **Moscow**<br>**Regional**<br>**Research**<br>**Clinical**<br>**Institute** |
| Tomograph model | Siemens Avanto 1.5T или Vario 3T | Philips Achieva 1.5T | Philips Achieva 1.5 T | Siemens Magnetom Aera 1.5 T |
| Basic parameters | Respiratory synchronization against the background of free breathing | Respiratory synchronization against the background of free breathing, with suppression of the signal from fat | Respiratory synchronization against the background of free breathing, with suppression of the signal from fat | Respiratory synchronization against the background of the patient's free breathing and synchronization with the ECG |
| TI (inversion time) | 300 ms | 280 ms | 280 ms | via TI-Scout MR sequence, 290-390 ms |
| TR (repetition time) | 5.4 ms | 5.3 ms | 5.3 ms | corresponded to the R–R interval (from 600 to 1100 ms) |
| TE (echo time) | 2.3 ms | 2.1 ms | 2.1 ms | 2,44 ms |
| Permission | 1.25□×□1.25□×□2.5 mm | 1.4□×□1.4□×□1.4 mm | 1.3□×□1.3□×□4.0 mm | 1,25×1,25×2,5 mm |
| Pre-scan | < 7d | < 7d | < 48h | <48h |
| Post scan | 3-6 months | 30 days | 3-6 months | 3-4 months |
| File Format | .nrrd |  |  | .nrrd |
| Number of images used during training | 11 |  |  | 21 |

While preparing the dataset, the part of the data was selected where the atrial wall could be distinguished well enough; the total number of such images was 11. This resulted in 24 images of patients before and after ablation. Some of them were not suitable for manual segmentation because of difficulties in defining the wall boundary; therefore, 21 images were used. Examples of images from the database we collected and from the open one are presented on Figure 1.

**Figure 1.**
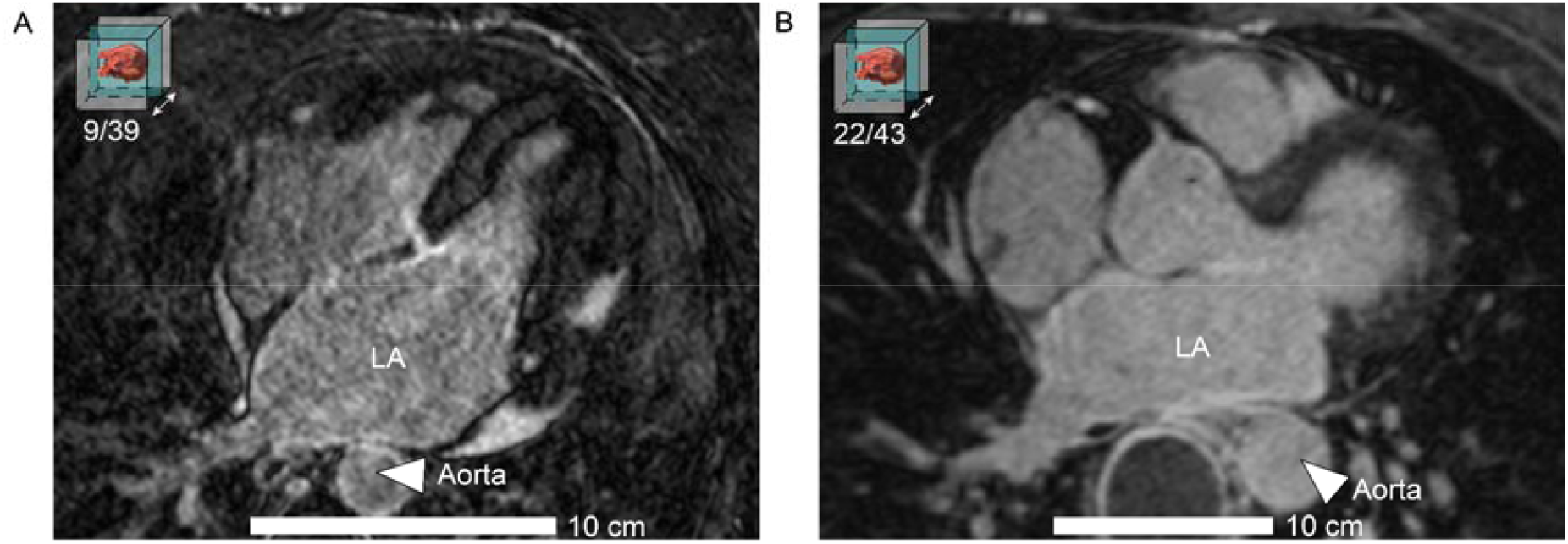
Original sagittal slices of MRI images of patients. A: Example of manually acquired data. B: Example of data from the open dataset cDEMRIS [12]. LA - left atrium, Aorta - aorta. A scale bar is shown in the lower part, the size corresponds to 10 cm. The slice number of the MRI image is shown in the upper left corner.

It has been shown that the interpretation of fibrotic foci on MRI can vary depending on the specialist who reads the images [11] and the center where they were obtained. Therefore, in order to standardize the process of MRI image segmentation and reduce the heterogeneity of the data, it is necessary to develop a single instruction for the segmentation technician, which will fix all the principles of image interpretation and make the segmentation sampling from different sources homogeneous.

### Construction of neural networks

Building UNet-like neural networks is a common approach to solving fMRI image segmentation problems [15], [16], we used such an architecture. Figure 2 shows a schematic of how the neural network works and our segmentation experiments with it.

**Figure 2.**
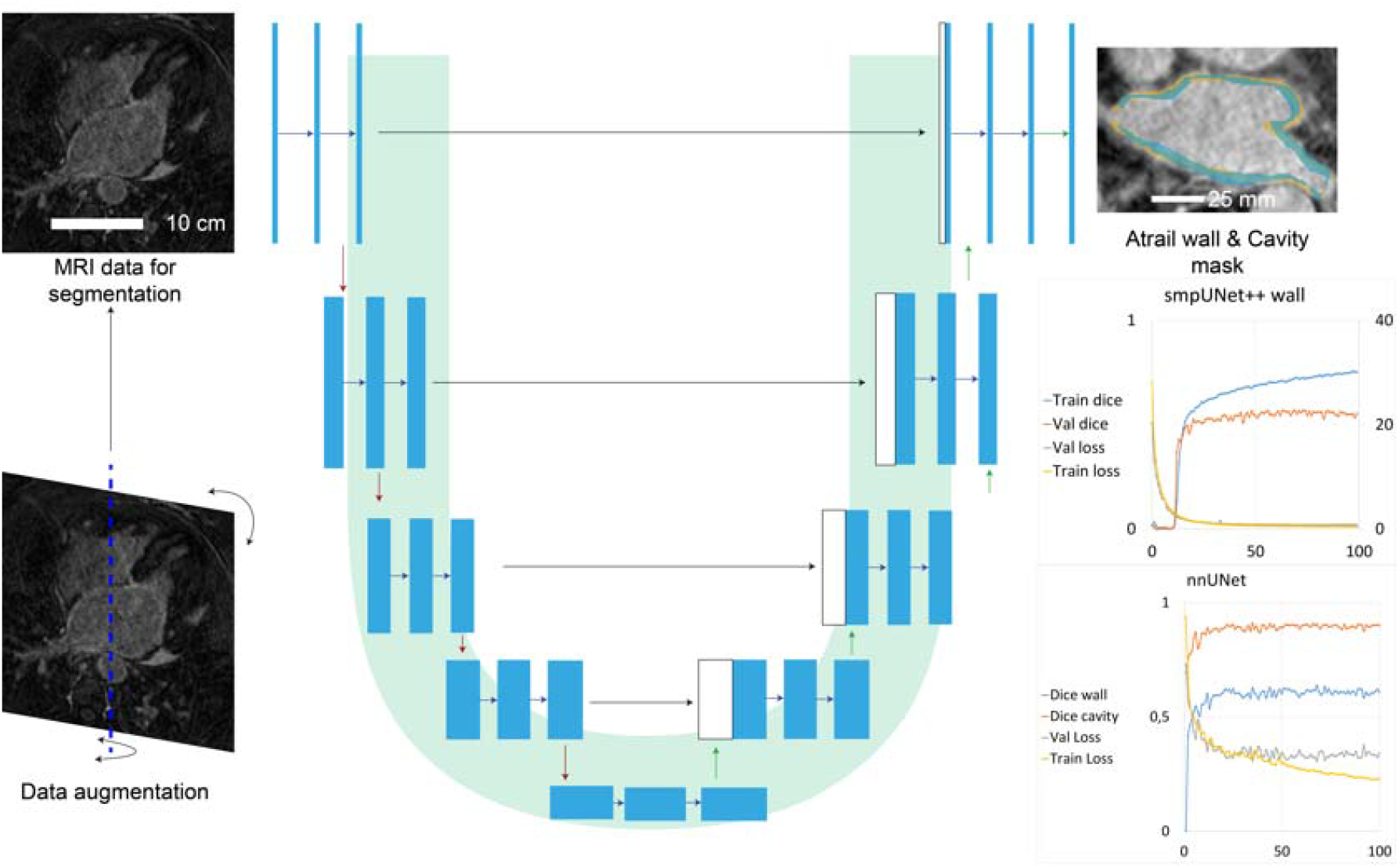
Scheme of the UNet-like neural network. Clockwise from bottom left: first, the segmented data are enhanced using standard methods, then the UNet-like neural network performs image segmentation and returns masks for the atrial cavity (blood pool) and atrial walls.

This paper presents the results of experiments on 2 types of neural networks: smpUNet ++ network [17] and nnUNet network [18]. The graphs in the lower right part of the figure show the results of training both types of networks, where for smpUNet++ network the training was performed only for wall segmentation, and for nnUNet network - for wall and cavity segmentation simultaneously (Dice [19] was used as metric, the error function was composite and consisted of two components BCE + DiceLoss [20], [12]).

Blue arrows in the figure are convolution operations - piecewise multiplication by a sliding matrix with a 3×3 kernel and ReLu activation function to add nonlinearity and model more complex - nonlinear dependencies in the data. Gray arrows are the concatenation of feature maps from different parts of the neural network, which is necessary to reduce the problem of vanishing gradients and also allows to transfer information from the left part - encoder to the right part - decoder. Red arrows are subsampling with a 2×2 kernel to reduce the size of the feature map and increase computational efficiency. Green arrows are upsampling with a 2×2 kernel to increase the feature map size. The last arrow is a convolution with a 1×1 kernel.

The architecture consists of an encoder - a constricting path for context capture and a decoder - a symmetric expanding path that allows for accurate localization of objects of interest. During training, single-channel fMRI images were fed to the input of the neural network. The output from the last layer was a mask - an image, each pixel of which belonged to one of two classes - atrial wall or background (atrial cavity / background for atrial cavity detection, respectively).

To reconstruct a 3-dimensional model of the atrium convenient for physician interpretation requires translating a set of atrial wall masks into a smooth structure (model) given the spatial resolution of the images and the distance between images. Recently, machine learning approaches have shown high performance [21]. RIFE [22] and FILM [23] neural networks were used for interpolation, the former showing better results (described in detail in Results).

Figure 3 shows a schematic diagram of the process of building a 3-dimensional model of the atrium from the patient’s MRI data. First, the training data set “Train MRI image set” is segmented with or without protocol, after which the neural network is trained, namely nnUNet [18] or smpUNet++ [17] architecture. A test sample “test MRI image” is used to evaluate the accuracy of the network. Then, based on the results of the atrial wall segmentation by the “Segmented mask” neural network, an atrial model is built as its interpolation. The white rectangle indicates the scale bar for the atrium, its size corresponds to 25 mm. In the interpolated atrial diagram, the mitral valve (MV), right inferior pulmonary vein (RIPV), and right superior pulmonary vein (RSPV) are indicated. Figure 3 also shows the results of training the neural networks with and without the use of the protocol labeled data. As can be seen, the use of the protocol significantly improves the training results: the accuracy of the atrial wall segmentation increased from Dice = 0.54 to Dice = 0.61 when the protocol was used for the nnUNet architecture [18].

**Figure 3.**
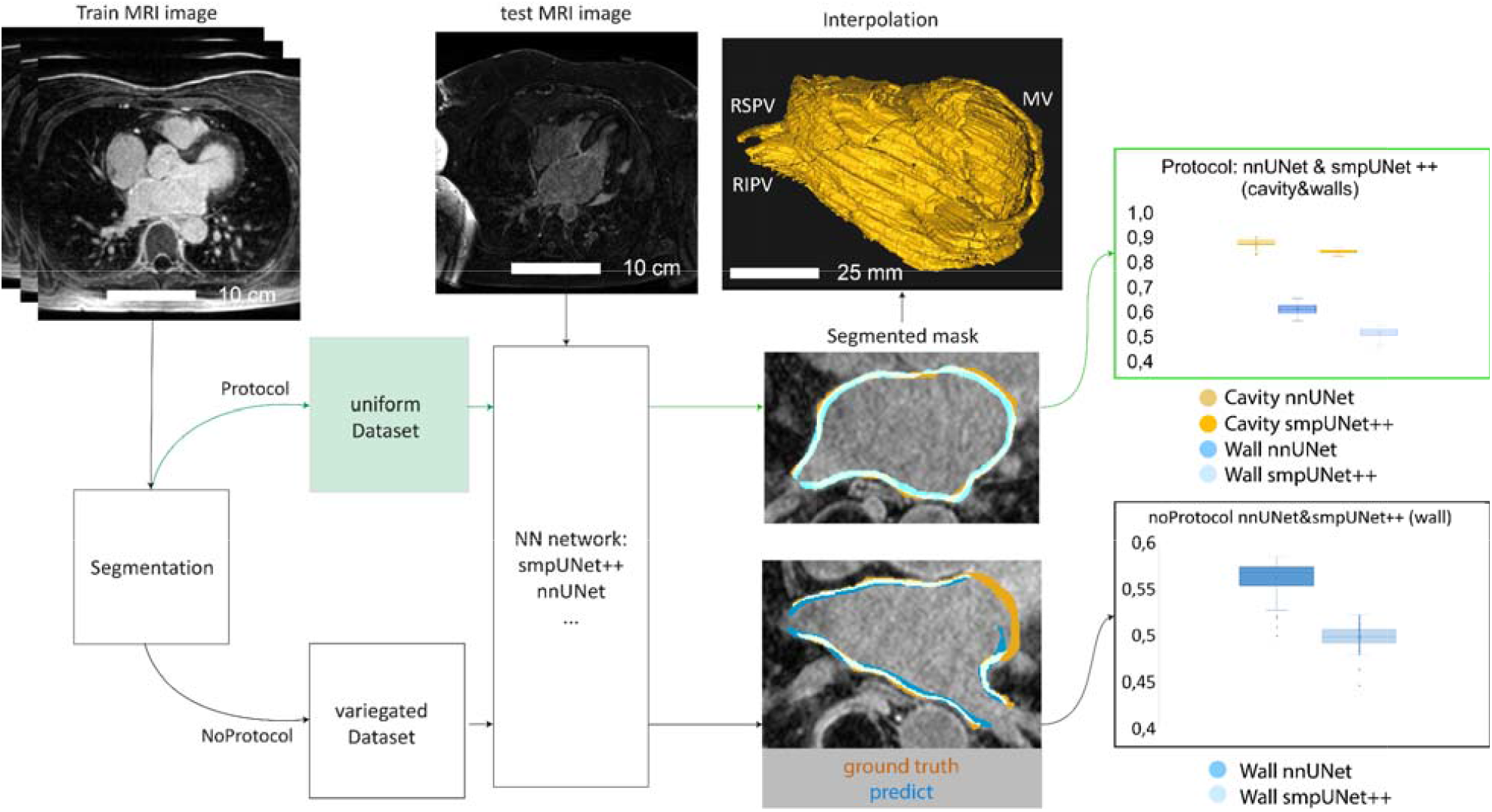
Schematic of how the 3-dimensional atrial model is generated from a patient’s MRI data: from a train MRI image set through various segmentation paths with or without protocol to a complete atrial model using interpolation. The white rectangle for the atrial model indicates a scale bar of 25 mm. In the interpolated atrial model, the mitral valve (MV), right inferior pulmonary vein (RIPV), and right superior pulmonary vein (RSPV) are indicated. The right part of the figure shows the results of the neural network training with and without the use of the protocol labeled data.

A wide range of experiments on training UNet-like neural networks, including: experiments on training Unet from scratch, architectures from the Torch Segmentation Models (smp) library [16]: smpUnet, smpUnet++, smpMANet, smpPSPNet, smpLinkNet, smpPAN, smpDeepLabV3, smpDeepLabV3+, smpFPN, pre-trained on the Imagenet dataset [19], 2 model architectures were selected for training: nnUNet [18] and smpUNet++ [17].

All models were implemented using the PyTorch framework [24] and trained using the Adam optimizer [25]. A value of 0.0001 was chosen for the learning rate factor, a hyperparameter that determines the order in which weights are adjusted to account for the gradient descent loss function. In all experiments, we ran 100 epochs of model training on an NVIDIA RTX A5000 GPU.

The Slicer framework [26] was used for manual segmentation of MRI images, and the work was performed independently by 2 physicians. The segmentation protocol was developed and agreed upon in advance, described in the corresponding section.

## Results

### 1. Development of criteria for the suitability of MRI images for segmentation

In order to segmentate each image and prepare the database, it was necessary to ensure that the criteria of its suitability for the analysis of the atrial wall and gadolinium accumulation in it were met. As a result of image selection, the following criteria were formulated for the suitability of MRI images for labeling:

1. If the wall is indistinguishable on any slice of the image, the image is considered unsuitable;
2. If a region of the atrial wall is indistinguishable on less than 3 slices, the region is reconstructed (approximately) from neighboring slices;
3. If the wall region is indistinguishable on 3 or more images, the image is considered unsuitable;
4. The edges of the pulmonary veins are segmented 2 cm from the base (approximately), with the scale bar displayed at the bottom of the screen;
5. The mitral valve remains open during segmentation (not considered part of the wall);
6. If more than 20% (visually) of the wall is artifactual (overlit), the image is considered inadequate.
7. Over-illumination is the excessive brightness of certain areas of the image caused by the accumulation of contrast (gadolinium) where it should not normally accumulate.

If all suitability criteria are met, the image can be segmented in the sequence described in Figure 4 and paragraph 2 of the results.

**Figure 4.**
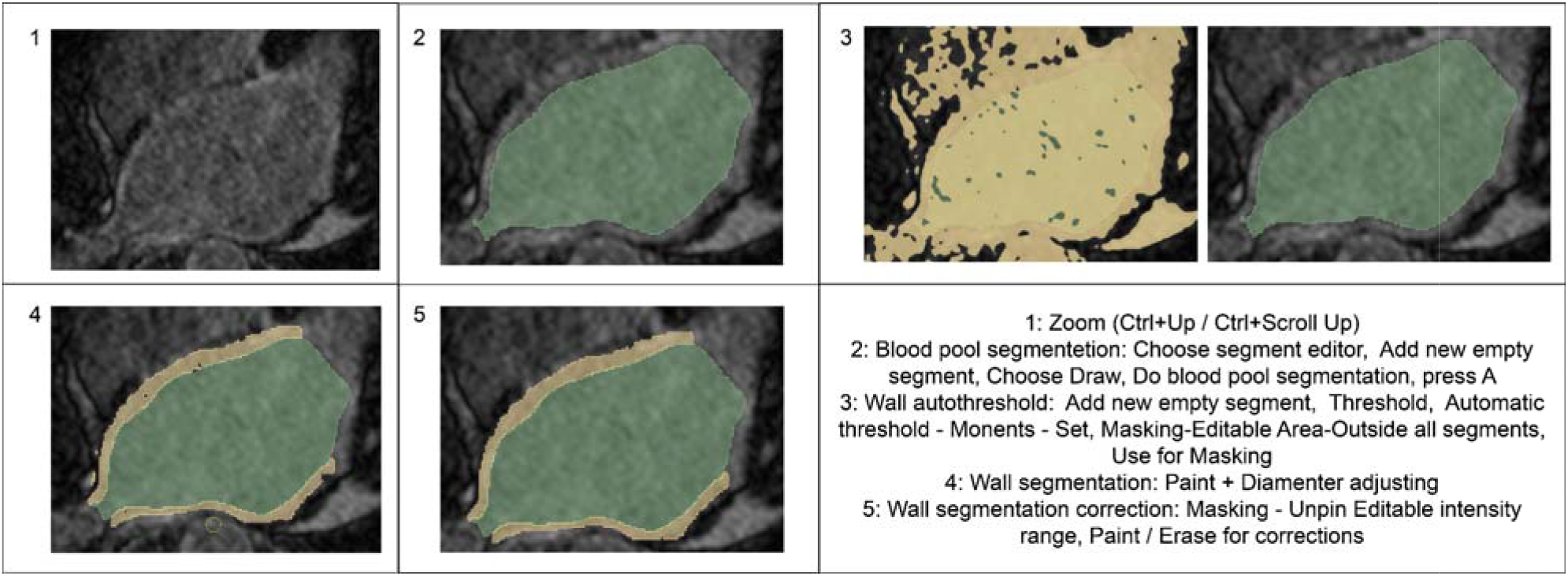
Illustration of the MRI image preprocessing protocol. Fields 1 - 5 show the panel view of working with MRI image slices while performing the corresponding segmentation step. The steps themselves are described (according to their numbers) in the lower right panel with instructions on how to use the Slicer interface tools.

### 2. Developing a universal atrial MRI image segmentation protocol for image preprocessing

To eliminate heterogeneity in image segmentation and to formalize the database preprocessing for neural network training, a universal protocol for atrial MRI image segmentation in Slicer was developed. The basic steps of blood pool and atrial wall segmentation in the images for network training are presented here and on Figure 4:

1. The image is enlarged on the screen so that it can be distinguished with pixel accuracy.
2. The patient’s blood pool is extracted from the image (pixel accurate) and a mask is assigned to this area (in the 3D Slicer interface [29]). The necessity of the mask is due to the fact that the walls will not overlap the blood pool during further slicing. The mask also makes the inner boundary more accurate and can also be used as a separate mask).
3. Using autoThreshold, i.e. Iso Data or Triangle method, an area close to the wall area is selected. The area is selected based on the contrast of each region in the image.
4. The wall is segmented with a brush of arbitrary thickness. Only the area selected in step 3 is automatically selected, which allows to speed up the wall segmentation, leaving less work for the next step (wall shape correction).
5. Errors in segmentation are corrected with a brush or an eraser. The final edge of the wall should be as flat as possible. There should be no holes or gaps in the wall thickness, the border of the blood pool should coincide with the inner border of the wall.
6. As a result of segmentation, files are saved with permissions: .nrrd - original image .seg.nrrd - segmented image

### 3. Training on partitioned data prior to developing a universal image preprocessing protocol

Prior to the segmentation standard protocol development, 32 images were segmented and a series of experiments were performed with a range of typical architectures, variations in augmentation approaches and loss functions. In this case, segmentation was performed using the Slicer program, but without defining precise universal steps to isolate wall, blood pull and fibrosis.

At the same time, the performance of the metrics using different architectures of neural networks remained quite low (Table 2), which can be attributed to the high heterogeneity of the segmented without protocol data. Therefore it was decided to develop a single protocol for atrial wall segmentation on MRI images. A summary of the experimental results of the atrial wall segmentation by 2 different neural networks, training on the data, which was founded before the protocol was developed is presented below on Table 2.

**Table 2.** A summary of the experimental results of the atrial wall segmentation by neural networks, training on segmented data without protocol.

| Model | Loss function | Dice |
| --- | --- | --- |
| smpUnet++ | DiceLoss+BCELoss | 0.56 |
| nnUnet - 3Dfullres, 1 fold | DiceLoss+BCELoss | 0.42 |

Thus, before the introduction of a common standard protocol for training dataset generation, a significant limitation affecting the quality of neural network performance was found. The manually segmented training dataset has a strong heterogeneity. Although this has been given special attention and a partitioning protocol has been specifically designed, the interpretation of such data, even with instruction, will differ to some extent. This problem has also been noted elsewhere [11]. As a consequence, we tested the quality of manual segmentation. For this purpose, we used 2 protocols: segmentation of the same image by different physicians and segmentation of the same image by the same physician but after a long time. Figure 5 shows an example comparing the segmentation of the same image by two physicians. It can be seen that the segmentation varies greatly, the Dice for it for some images may be lower values than when using the trained model. It was found that in fact, the specified Dice value for manual segmentation is the upper limit of the metric and for the neural network, it is not possible to surpass it when using a training dataset without a protocol.

**Figure 5:**
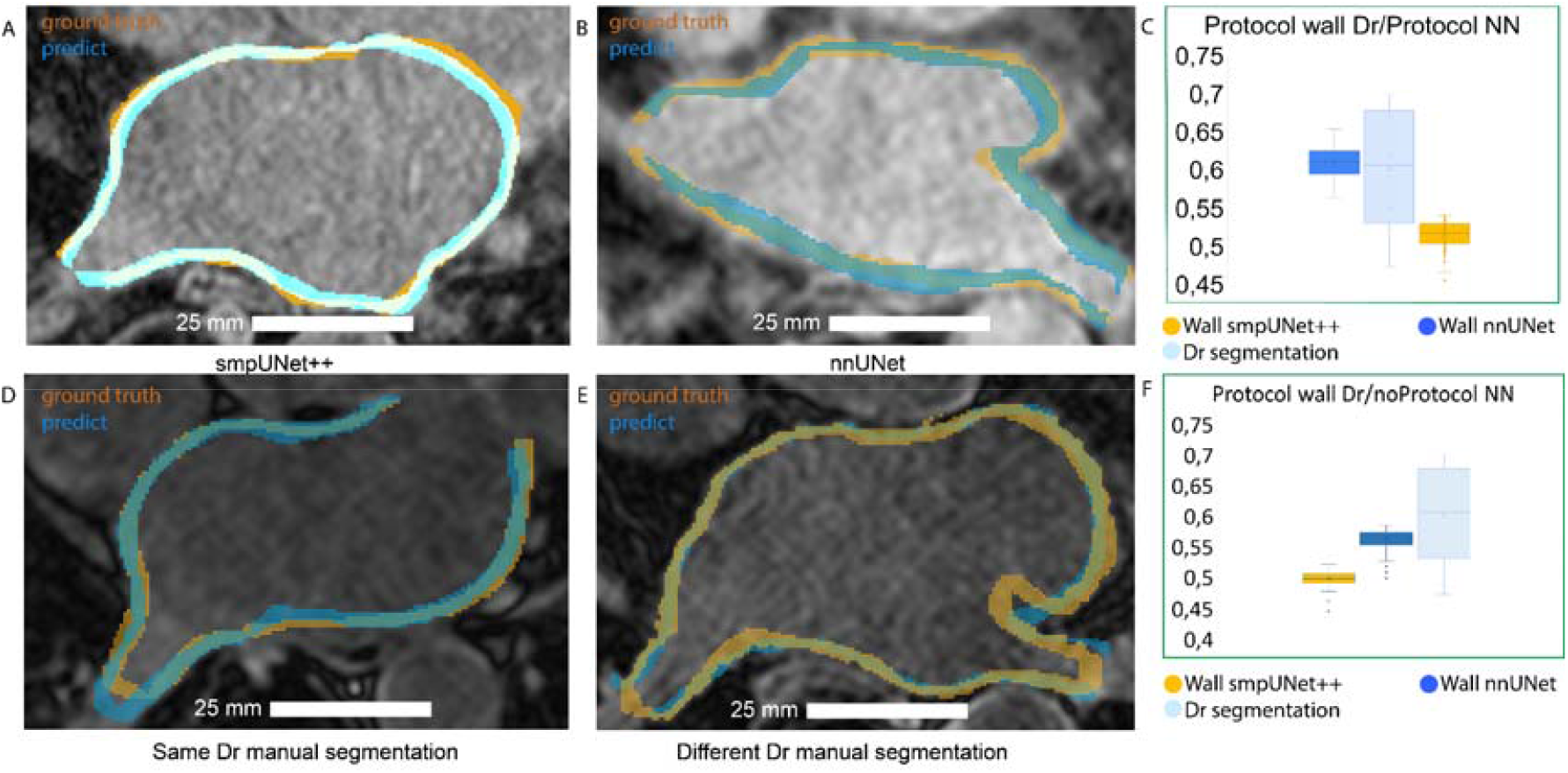
Analysis of the homogeneity of manual markup: it can be seen that the markup of the same images differs from one doctor to another and when repeatedly marked up by the same doctor. The corresponding Dice value is a natural constraint from above for the results of the neural network. A: the illustration of the operation of smpUNet++; B: the illustration of the work of nnUNet; C: the comparison of the physician and neural networks’ accuracy of segmentation to the wall using the protocol. For the physician Dice was 0.6 +-0.08 whereas for the neural network it was 0.64; D: the comparison of the markup of the same image taken by the same marker; E: the comparison of the segmentation of the same image made by different markers; F: the comparison of the accuracy of the markup of the physician and neural networks per wall without using the protocol. For the physician Dice was 0.6+-0.08, whereas for the neural network it was 0.64.

### 4. Training on data segmented using a standard protocol

Due to the lack of a standard protocol, the quality of the neural networks did not change as the training data set increased. Therefore, we developed a protocol for manual segmentation and preprocessing of training datasets, as described in point 2 above. The dice parameter in Table 3 shows that the protocol significantly improved the quality of trained neural network segmentation. If we compare manual segmentation and segmentation using a trained neural network, the differences are minimal. Examples of comparing the performance of the neural network and manual segmentation using the protocol are shown in the following figure 6.

**Table 3.** A summary of the experimental results of the atrial wall segmentation by neural networks, training on segmented data with standard protocol.

| Model | Loss function | Target | Dice |
| --- | --- | --- | --- |
| smpUNet++ | DiceLoss+BCELoss | walls | 0.64 |
| smpUNet++ | DiceLoss+BCELoss | pull | 0.9 |
| nnUnet - 3Dfullres, 1 fold | DiceLoss+BCELoss | walls+pull | 0.59 and 0.87 |

**Figure 6.**
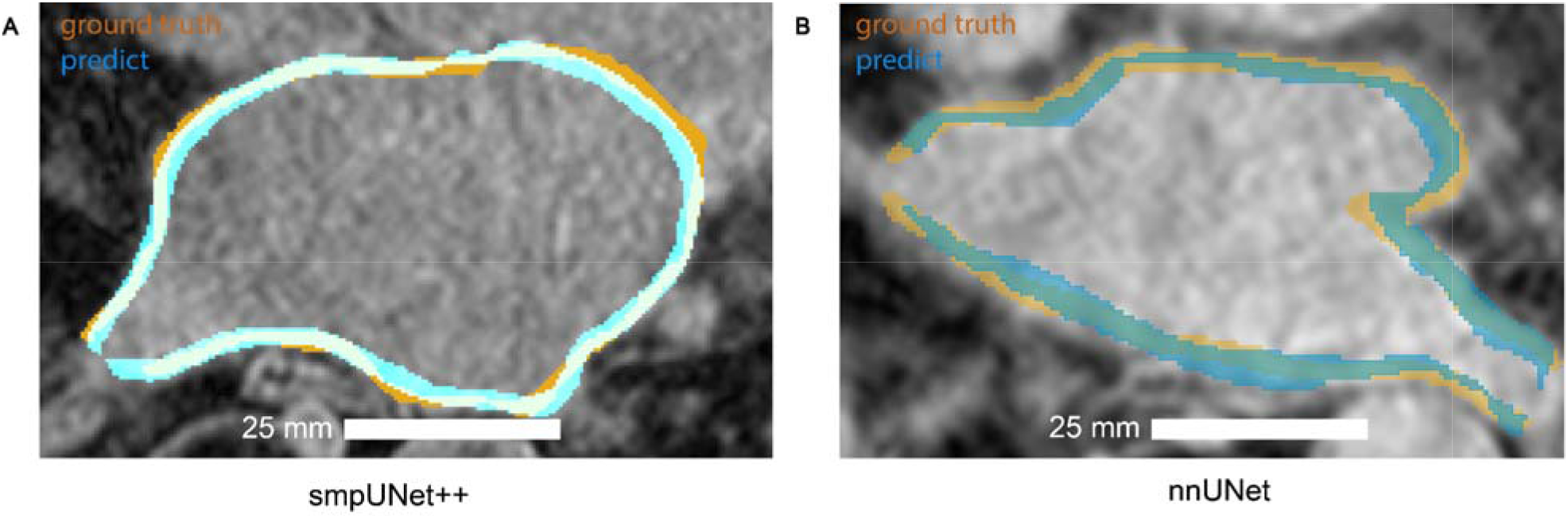
Illustration of neural networks trained with smpUNet++ (A) and nnUNet (B). Blue is the result of the network segmentation (prediction), orange is the physician’s manual marking (ground truth).

Thus, the results of training on data partitioned using the protocol are superior to the results of training on data partitioned without the protocol. This may be due to the fact that the data has become more homogeneous and the quality of partitioning has generally increased. Moreover, there is a correlation that the limit of efficiency of segmentation with the help of neural network in this situation does not depend on the amount of data for training, but on the use of a single protocol for partitioning the training data set. The application of the protocol had a positive effect on the training results, it can be considered reasonable.

### 5. Interpolation of segmented MRI images to create a 3D model of the atrium

A number of experiments were conducted to select the best method of image interpolation. Since the resolution of the used images ranges from 1.5 to 0.625 mm per pixel and the distance between slices is more than 2 mm, it is necessary to interpolate the images along the vertical axis in order to obtain a homogeneous grid. Classical methods such as linear interpolation, for example, cannot be used correctly in this case, because in this case the heart surface will not have a physiological shape.

Instead of classical methods, interpolation of the images was performed using machine learning methods to select the optimal interpolation method. For this purpose, the models used to increase the capture rate (fps) of video recordings were used. The following models were considered:

1. FILM: Frame Interpolation for Large Motion
2. RIFE: Real-Time Intermediate Flow Estimation for Video Frame Interpolation

All experiments had a quantitative evaluation of the performance of the interpolation models. The peak signal-to-noise ratio (PSNR) metric was chosen as the main metric:

The RIFE algorithm showed better results compared to FILM on selected example images, as shown in the examples in Table 4.

**Table 4.** Comparison of the results of interpolation models using the PSNR metric using several patient examples.

| Patient image number | Google (FILM) | RIFE v3.x HD model |
| --- | --- | --- |
| 11 | 27.99 | 28.07 |
| 14 | 27.4 | 27.6 |
| 20 | 26.6 | 26.7 |

According to the results of the comparison of metrics, the RIFE model was chosen, which demonstrated higher results. It can be seen that the difference for the problem to be solved is not significant. As an example, Figure 7 shows an illustration of atrial wall interpolation by RIFE and FILM models. It can be seen that the interpolation in Figure 7A is much more convenient than the visual one for interpretation, although the difference in metrics is small.

**Figure 7.**
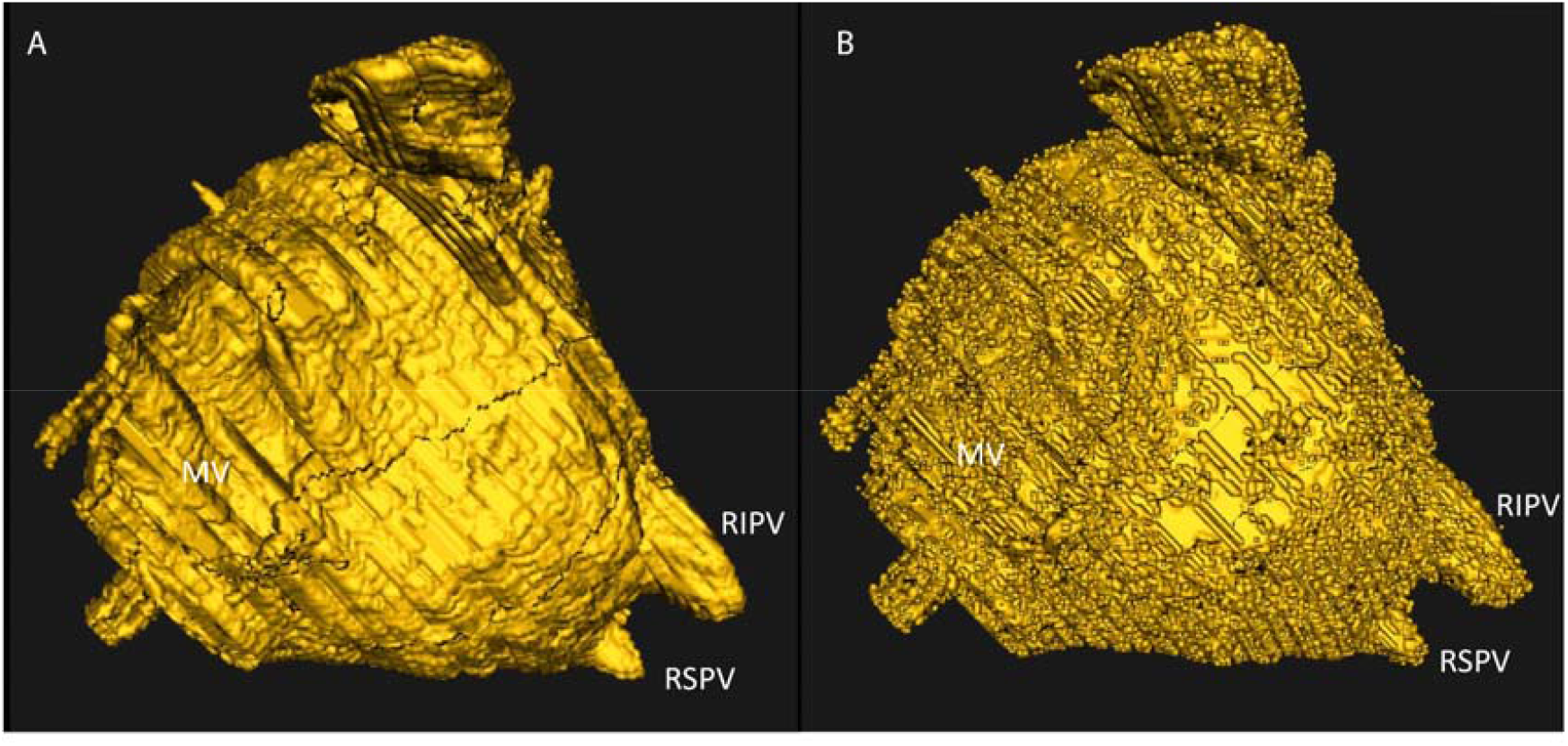
Examples of image interpolation using the RIFE (A) and FILM (B) neural network. The diagram of the interpolated atrium shows: mitral valve (MV), right inferior vena cava (RIPV), right superior vena cava (RSPV).

## Discussion

The task of left atrial wall segmentation and fibrosis, as well as 3D reconstruction of human atria, is currently of great clinical importance [8]. Meanwhile, the current routine manual segmentation of atria on medical images is a time-consuming and error-prone process. Our work demonstrates, among other things, how much the markups of several physicians can differ (Figure 5).

One of the solutions to the wall segmentation problem at the moment is the application of machine learning. In the course of solving the problem, deep learning approaches were developed for atrial segmentation, which achieved high accuracy (>90% of Dice score) [27]. But in this segment of development, the problems of significant divergence from other manual segmentations, problems of optimal deep learning architecture that would ensure robustness while achieving the best performance still remain [11]. In this paper, we have tried to focus on the model training problem by taking the most popular architectures in solving U-Net series MRI segmentation problems [27]. A problem with many studies of this kind is the preprocessing of the data for training: for example, if the data is acquired from a single MRI machine or labeled by a single person [27; 28]. In the presented study, we show that the only optimal method to improve accuracy is to standardize the protocol for preprocessing the training data set. In a general sense, there are no standards for manual segmentation, making it nearly impossible to train neural networks on this data. After introducing the segmentation protocol, practicing it with several independent clinicians, and preprocessing the data for training with it, we noticed an improvement in the quality of the neural networks. Dice reached about 0.9 (Table 3). The introduced manual segmentation protocol can be improved in the future, which will naturally produce data of higher homogeneity.

After the introduction of the protocol, the training revealed some more fundamental errors in the performance of the trained models, illustrated in Figure 8. These are errors such as:

**Figure 8.**
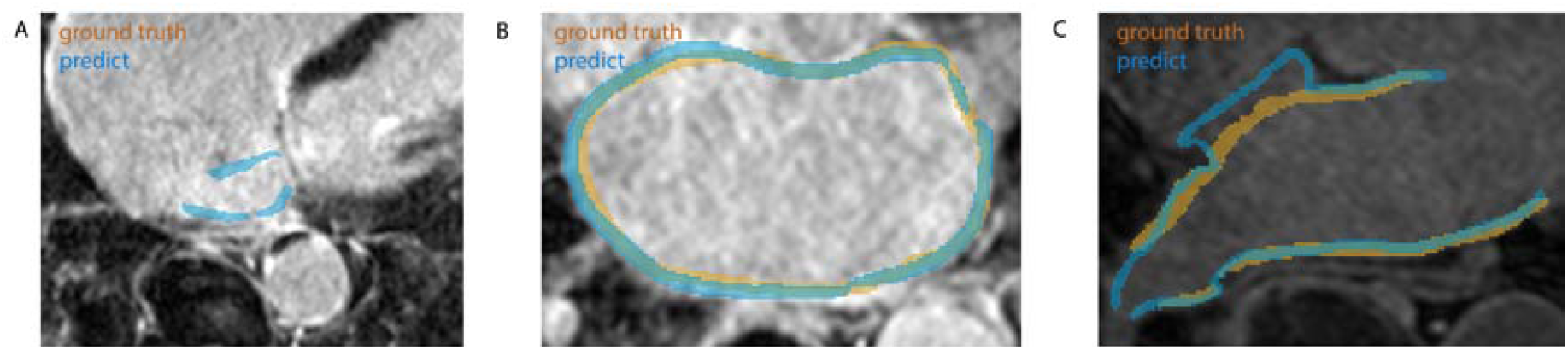
Illustration of typical errors occurring during automatic atrial wall segmentation. A: The model segments regions on slices that do not have atrial wall (second-order error); B: The model does not find the entire wall (second-order error on heart regions); C: The model selects individual heart regions other than atria (first-order error on heart tissue). Blue indicates model prediction, yellow indicates manual segmentation by the physician.

A. The model segments regions on slices where there is no atrial wall. The fact is that atrial walls on images fundamentally look the same as ventricular walls and are chosen by the physician mainly for anatomical reasons;
B. The model does not find the entire wall. Difficulty in interpretation may be due to the fact that on some images the difference between the brightness of the atrial wall and the inner cavity is not very large;
C. The model highlights separate regions of the heart other than the atrium. This may arise because the brightness is higher not only at the atrial wall but also in other areas of the image. We expect that increasing the size of the dataset will overcome this problem.

Speaking about the limitations of the presented study, it is worth noting the small amount of data. The maximum number of data in the training sample was 32 images, 11 of which were taken from the open dataset cDEMRIS. In principle, unfortunately, there is a database problem of segmented MRI image databases, which has been shown in several other works [29]. Another prerequisite for the limitation of segmentation quality may be the small area of the object in the image - as a rule it occupies less than 20% of the whole image area. It is possible to approach the solution of this problem by preprocessing the images with limitation of the analyzed area. In our case, the images were cropped, which increased the preset efficiency. In the future, it is possible to restrict the analysis area more strongly by using reference points, for example, to define on the image the boundaries of the thorax and analyze only in the inner region. It should be mentioned that a natural limitation when analyzing images is their quality. The resolution of the wall in the images requires a sufficiently high quality resolution of the images themselves to capture the shape of the wall. In addition, it is important to effectively stain the tissue with contrast (gadolinium) by taking images at the optimal moment.

In addition to testing U-NET architectures, this paper presents a comparison of architectures for interpolation and construction of 3D atrial models. This construction is a final point for physicians, a result that can already be directly handled in navigation operating systems. This work is part of the creation of a cardiac ablation correction system. Other important parts of such a system are a tool for performing wave dynamics simulations in cardiac tissue and a tool for cell array simulation. Once all parts of the system are integrated with each other, it will be tested on clinical cases.

Suggested potential limitations of the metrics values in our case could be:

1. Insufficient amount of data for this task;
2. Small area of the segmentation object;
3. Selection of an inappropriate protocol for data partitioning;
4. Too much noise in the data;
5. Inappropriate data preprocessing.

Note that often better results are obtained using data from a single machine and using a single partitioner, which automatically increases the homogeneity of the data, but does not provide any information about the algorithm’s performance on data from other sources and is more likely to lead to errors at the MRI image analysis stage.

In addition to rotation and offset augmentation, CenterCrop was also used to resize to 320×320 pixels, but this increased overtraining without improving the metric. Conventional contrast equalization made the images overexposed, and CLAHE adaptive contrast equalization [30] had no effect on the metric. Increasing the number of channels from one to three by concatenation to apply encoders trained on three-channel images from the Imagenet dataset did not change the metric. Calculating the mean and standard deviation on the training portion of the dataset to further normalize the data also did not increase the metric.

In the future, it is planned to obtain and partition more data and conduct experiments on training models on an extended dataset partitioned using the protocols [31]. In addition, it is planned to continue experimenting with data preprocessing and do post-processing of the results obtained from the model. Post-processing could improve the quality of segmentation by eliminating regions outside the atrium that are falsely detected by the model, for example, by identifying the region where the atrium is located and zeroing out the surrounding segmentation results some of the errors could be eliminated.

## Conclusions

In this paper, we were able to compare the training results of U-NET architecture neural networks depending on the training data set. Based on these data, a protocol for preprocessing MRI images was developed. It was shown that such a protocol significantly increases the Dice after training the neural network. The study also collected a proprietary database of manually segmented images. In addition, we compared neural networks for interpolation of segmented images based on the training results.

## Data Availability

The database announced in the work can be provided upon request using the link: https://doi.org/10.5281/zenodo.11102309
The machine learning code presented in the work can be found by the link: https://github.com/CardioBioLab/MRI_segmentation_paper

## Supplementary Materials

Appendix 2 “Full results of training neural network models on data segmented without a protocol”

Appendix 1 “Illustrated Image Segmentation Protocol”

## Author Contributions

Conceptualization, B.A., S.V., Ts.V. and S.M.; methodology, B.A., S.V., Ts.V., K.V., S.M.; software, B.A., K.A., P.D., S.A., D.A., Z.A.; validation, B.A., K.A., P.D., S.A., D.A., Z.A., Sh.R., A.A.; formal analysis, B.A., K.A., P.D., S.A., D.A., Z.A., Sh.R., A.A.; investigation, B.A., K.A., P.D., S.A., D.A., Z.A., Sh.R., A.A., S.M., K.V., S.V., Ts.V.; resources, B.A., K.A., P.D., S.A., D.A., Z.A., Sh.R., A.A.; data curation, B.A., K.A., P.D., S.A., D.A., Z.A., Sh.R., A.A.; writing—original draft preparation, B.A., S.V., Ts.V., K.V., S.M.; writing—review and editing, B.A., S.V., Ts.V., K.V., S.M.; visualization, B.A., Ts.V., S.M.; supervision,B.A., S.V., Ts.V. and S.M.; project administration, B.A., S.V., Ts.V. and S.M.; funding acquisition, Ts.V.. All authors have read and agreed to the published version of the manuscript.

## Institutional Review Board Statement

The study was conducted in accordance with the Declaration of Helsinki, and approved by the Ethics Committee of M.F. Vladimirsky Moscow Regional Clinical Research Institute (protocol №7 from 18.04.2024).

## Informed Consent Statement

Informed consent was obtained from all subjects involved in the study.

## Data Availability Statement

The database announced in the work can be provided upon request using the link: https://doi.org/10.5281/zenodo.11102309 The machine learning code presented in the work can be found by the link: https://github.com/CardioBioLab/MRI_segmentation_paper

## Acknowledgments

The work was mainly supported by the Ministry of Science and Higher Education of the Russian Federation in the scope of the government assignment (Agreement 075-03-2023-106 13 January 2023) and M.F. Vladimirsky Moscow Regional Clinical Research Institute state grant #55. So, we would like to express special gratitude to the administration of the ITMO university and MIPT for the financial support of the authors. We would also like to express special gratitude to the administration of the Almetyevsk State Oil institute and «Tatneft» company for supporting the project.

### Appendix 1

Illustrated Image Segmentation Protocol:

1. The image is enlarged on the screen to the point where it can be distinguished with pixel accuracy
2. A pool of patient’s blood is selected (pixel-perfect) on the image, this area is assigned as a mask (in the 3D Slicer interface [29]). This is necessary to avoid overlapping the blood pool during further wall marking and to make the internal boundary more precise, and can also be used as a separate mask).

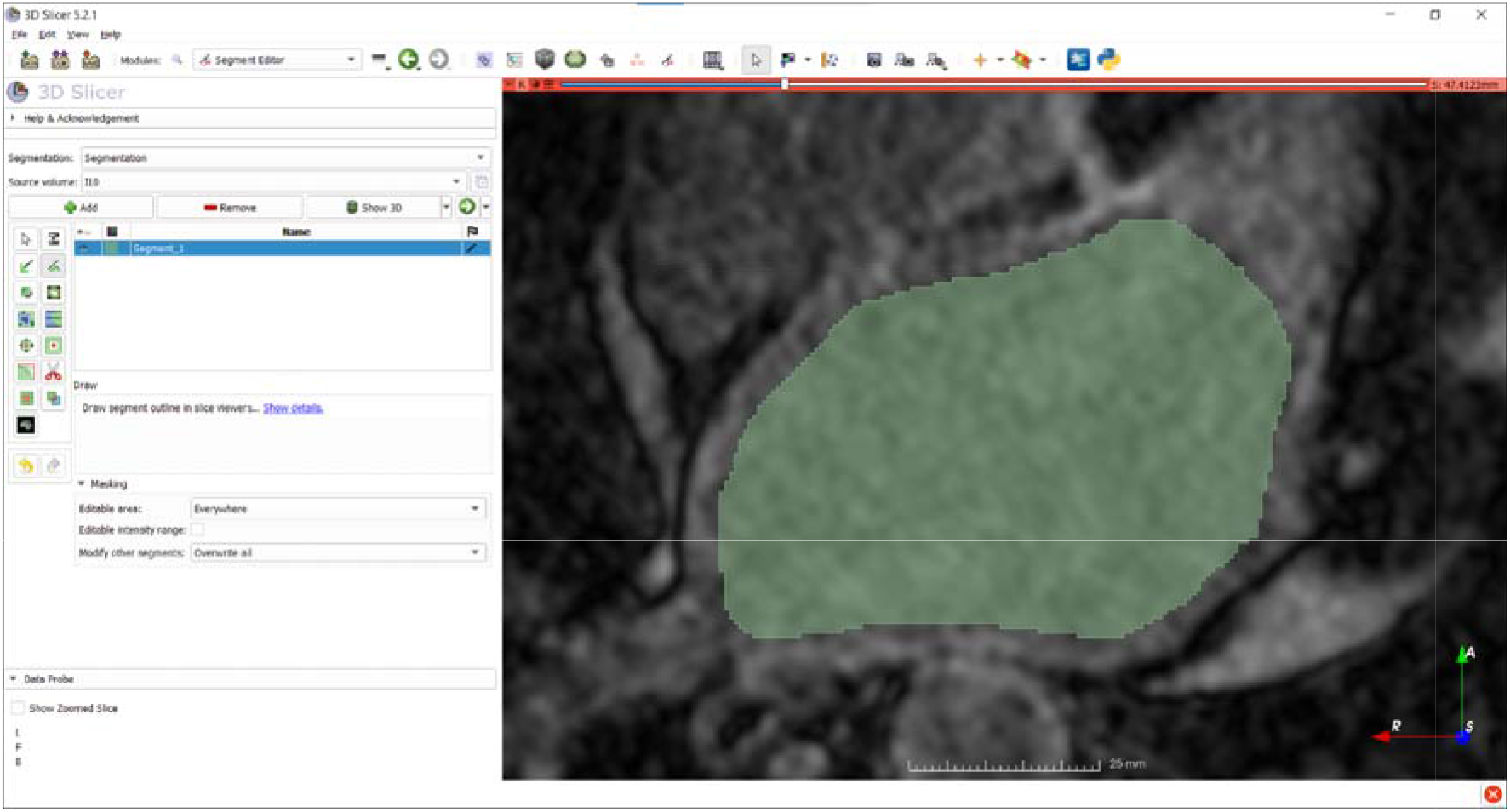
3. Using autoThreshold (Iso Data or Triangle method), a region close to the wall region is selected (to facilitate further partitioning). This is done based on the contrast of the individual regions in the image

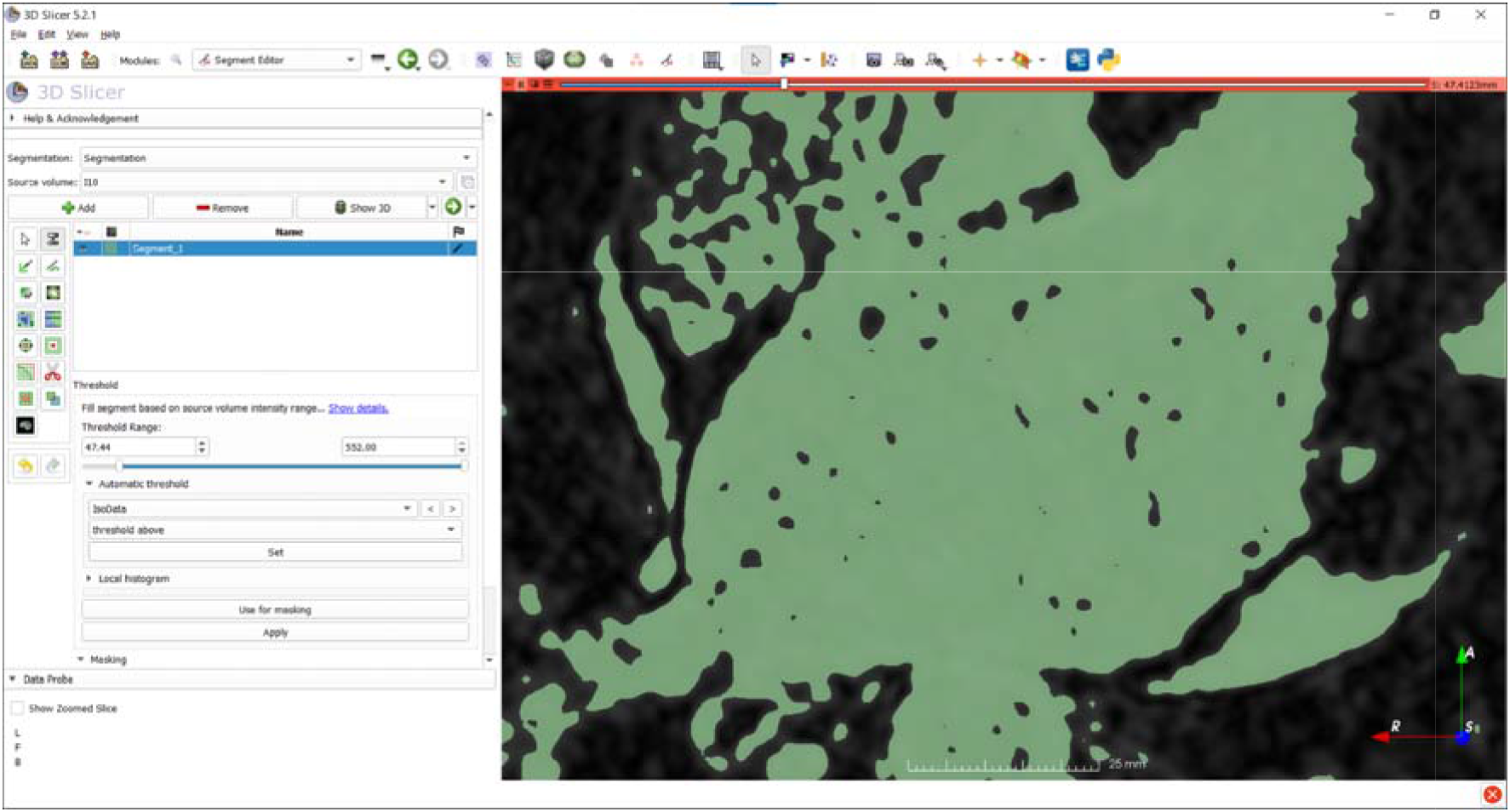
4. The wall is marked with a brush of any thickness (convenient for the marker). Only the area selected in step 3 is automatically selected. This allows to speed up wall marking, leaving less work for the next step (correcting the wall shape).

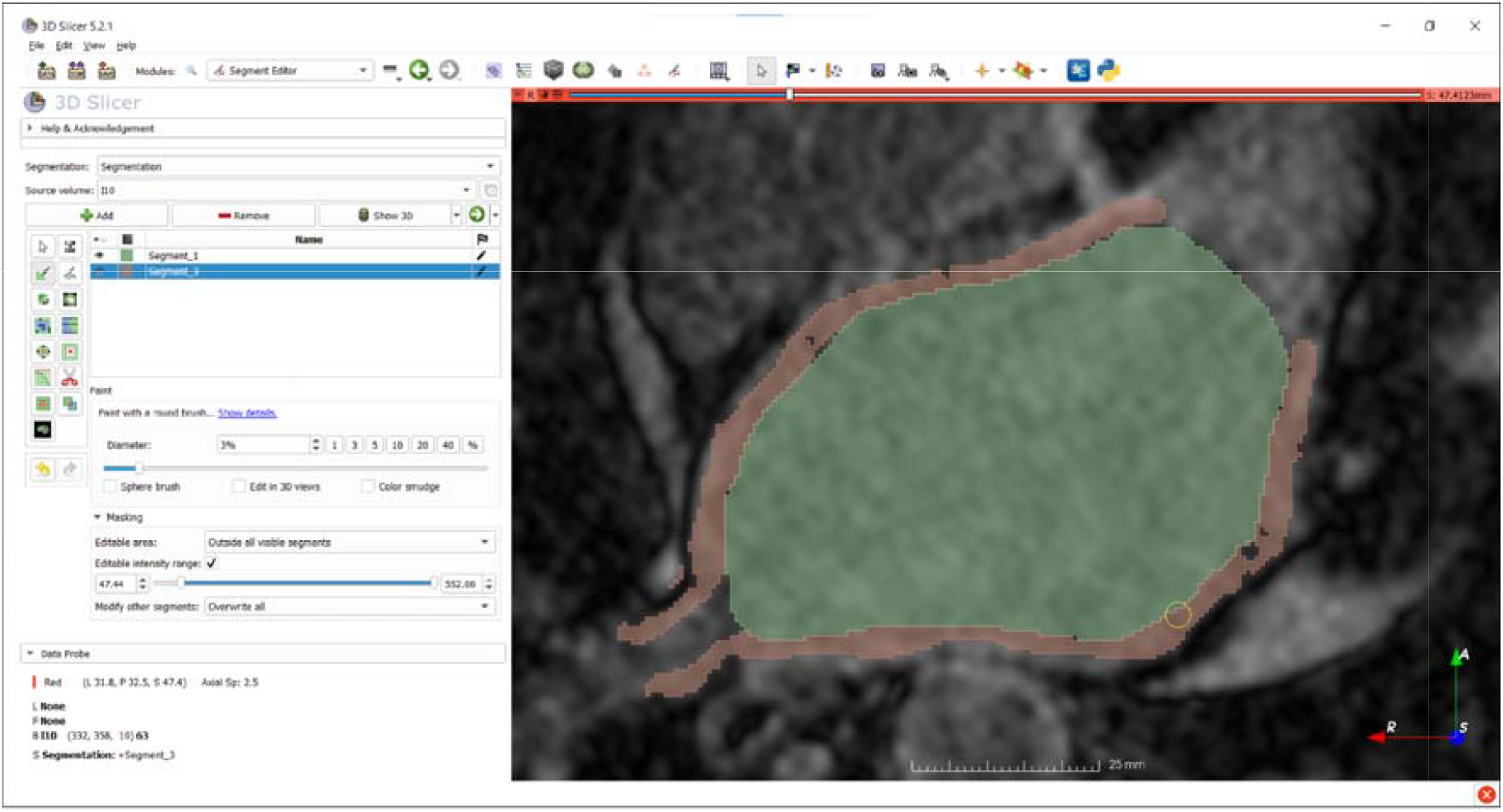
5. Errors in marking are corrected with a brush or eraser, the final edge of the wall should be as flat as possible. There should be no holes or gaps in the wall thickness, the border of the blood pool should coincide with the inner border of the wall

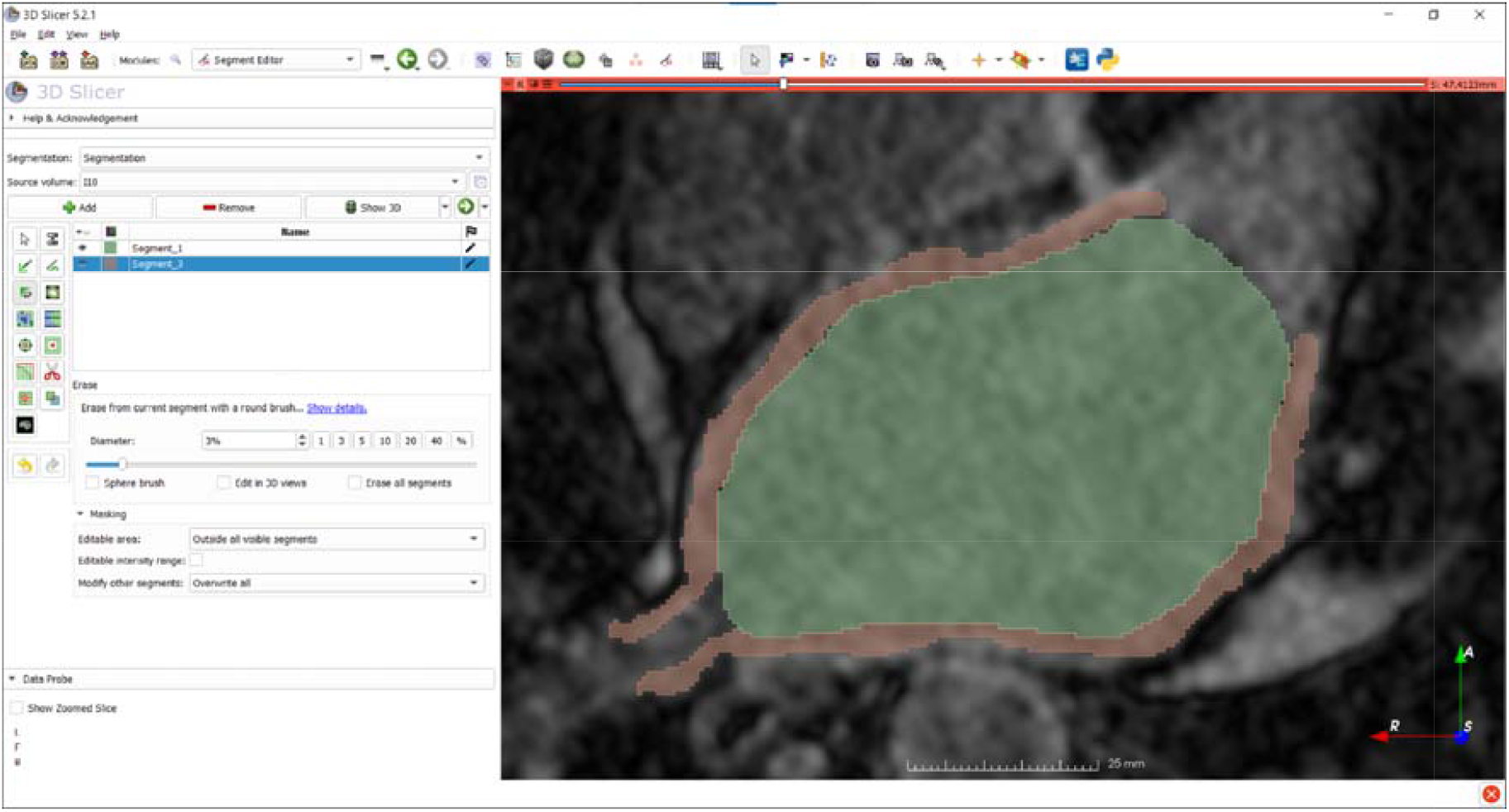

As a result of partitioning, files with permissions are saved:

1. .nrrd - original image
2. .seg.nrrd - segmented atrial wall and blood pool masks

### Appendix 2

Full results of training neural network models on data segmented without a protocol

| Model | Loss function | Dice |
| --- | --- | --- |
| Unet | DiceLoss+BCELoss | 0.40 |
| Unet++ | DiceLoss+BCELoss | 0.42 |
| Unet (brain MRI preprocessed) | BCELoss | 0.39 |
| smpUnet | BCELoss | 0.41 |
| smpUnet++ | BCELoss | 0.40 |
| smpMANet | DiceLoss+BCELoss | 0.43 |
| smpPSPNet | DiceLoss+BCELoss | 0.38 |
| smpLinkNet | DiceLoss+BCELoss | 0.44 |
| smpPAN | DiceLoss+BCELoss | 0.42 |
| smpDeepLabV3 | DiceLoss+BCELoss | 0.41 |
| smpDeepLabV3+ | DiceLoss+BCELoss | 0.43 |
| smpFPN | DiceLoss+BCELoss | 0.45 |

